# Vitamin D – contrary to vitamin K – does not associate with clinical outcome in hospitalized COVID-19 patients

**DOI:** 10.1101/2020.11.07.20227512

**Authors:** Jona Walk, Anton S.M. Dofferhoff, Jody M.W. van den Ouweland, Henny van Daal, Rob Janssen

**Affiliations:** Department of Internal Medicine, Medicine, Canisius-Wilhelmina Hospital, Nijmegen, The Netherlands; Department of Clinical Chemistry, Medicine, Canisius-Wilhelmina Hospital, Nijmegen, The Netherlands; Department of Pulmonary Medicine, Canisius-Wilhelmina Hospital, Nijmegen, The Netherlands

**Keywords:** COVID-19, desmosine, elastic fibers, vitamin D, vitamin K, 25-hydroxyvitamin D

## Abstract

SARS-CoV-2 causes remarkably variable disease from asymptomatic individuals to respiratory insufficiency and coagulopathy. Vitamin K deficiency was recently found to associate with clinical outcome in a cohort of COVID-19 patients. Vitamin D has been hypothesized to reduce disease susceptibility by modulating inflammation, yet little is known about its role in disease severity. Considering the critical interaction between vitamin K and vitamin D in calcium and elastic fiber metabolism, we determined vitamin D status in the same cohort of 135 hospitalized COVID-19 patients by measuring blood 25(OH)D levels. We found no difference in vitamin D status between those with good and poor outcome (defined as intubation and/or death). Instead, we found vitamin D sufficient persons (25(OH)D >50 nmol/L) had accelerated elastic fiber degradation compared to those with mild deficiency (25(OH)D 25-50 nmol/L). Based on these findings, we hypothesize that vitamin D might have both favorable anti-inflammatory and unfavorable pro-calcification effects during COVID-19 and that vitamin K might compensate for the latter.

## Introduction

Severe acute respiratory syndrome (SARS)-coronavirus (CoV)-2 is the causal agent of coronavirus disease 2019 (COVID-19). Interindividual variability of COVID-19 severity is remarkably high. The majority of infected individuals are asymptomatic or have mild to moderate symptoms. However, a significant minority develops life-threatening respiratory failure. The former group is mainly responsible for the rapid global spread of the virus, whereas the latter put pressure on medical resources and forced governments to proclaim lockdowns. This perfect storm unleashed a worldwide health and socioeconomic crisis of unprecedented proportions in modern times. Modifiable determinants of disease severity are important to identify as they have the potential to improve patient outcomes and reduce the need for restrictive measures.

We recently proposed vitamin K status as such a modulator [1] based on the strong association between vitamin K insufficiency and poor outcome in a cohort of COVID-19 patients [2]. Vitamin K is a fat-soluble vitamin essential for the activation of coagulation factors in the liver, as well as extrahepatic proteins including Matrix Gla Protein (MGP) [3]. MGP may be especially relevant to COVID-19. It is a potent inhibitor of soft tissue calcification and elastic fiber degradation [4, 5] with a protective role in dynamic tissues like lungs and arteries [6]. Accelerated elastic fiber degradation has been demonstrated in COVID-19 [2].

In recent months, a great deal of attention has been paid to the role of vitamin D in COVID-19 [7], with many suggesting that vitamin D administration might prevent infection or improve disease outcomes [8]. Vitamin D is mainly endogenously synthesized in the skin under the influence of sunlight and can be exogenously obtained from certain foods or supplements as well. It plays a modulatory role in both innate and adaptive immunity [9]. The potent anti-inflammatory properties of vitamin D might be expected to dampen SARS-CoV-2-related cytokine storm [10]. Indeed, administration of vitamin D was demonstrated to be protective against acute respiratory tract infection in a meta-analysis before the emergence of SARS-CoV-2 [11]. However, studies assessing the potential role of vitamin D in COVID-19 have had contradictory results [12, 13].

It is important to consider vitamin D supplementation in the context of vitamin K and elastic fiber metabolism as these pathways are interrelated [14]. Namely, high-dose vitamin D administration in rats has been shown to stimulate calcification and degradation of elastic fibers [15, 16]. Presumably as a compensatory mechanism, the MGP-gene promotor region contains a binding site that upregulates expression in response to vitamin D attachment [17]. For this effect to be protective, high levels of vitamin K are necessary to activate newly synthesized MGP. The effect of vitamin D on elastic fiber metabolism in COVID-19 patients has not yet been studied.

We hypothesized that vitamin D and K might be interrelated determinants of COVID-19 outcome through modulation of both the immune system and elastic fiber metabolism.

## Methods

### Subjects

Vitamin D status was determined in samples from a previously described patient cohort [2]. In short, 135 patients hospitalized for COVID-19 in the Canisius-Wilhelmina Hospital in March and April 2020 were included in the analysis. After SARS-CoV-2 infection was confirmed by Real Time polymerase chain-reaction testing, blood was sampled in EDTA tubes from each patient, centrifuged and plasma was frozen at −80 degrees Celsius. Patients were followed-up until: 1) discharge from the hospital, 2) intubation and invasive mechanical ventilation, or 3) death. Outcome of COVID-19 patients was classified as ‘good’ if they were discharged from the hospital without the need for invasive ventilation, and ‘poor’ if they required intubation and mechanical ventilation and/or died.

### 25-hydroxyvitamin D

Vitamin D was measured in EDTA plasma by quantifying 25-hydroxyvitamin D (25(OH)D) levels using a validated liquid chromatography-tandem mass spectrometry (LC-MS/MS) as previously described [18]. Circulating 25(OH) D is regarded as the best indicator of vitamin D status, because it reflects both production in the skin and dietary intake. For this study vitamin D sufficiency was defined as a 25(OH)D concentration greater than or equal to 50 nmol/L, insufficiency between 25 to 50 nmol/L and deficiency as less than 25 nmol/L.

### Desmosine

Desmosine and isodesmosine (DES) levels were determined in EDTA plasma and used as a marker for the rate of elastic fiber degradation. DES are formed during the cross-linking of tropo-elastin polymers and are released in the bloodstream after degradation of elastic fibers. Therefore, plasma (p)DES directly reflects the rate of systemic elastic fiber degradation. Both DES fractions were measured using LC-MS/MS as previously described [2]. Coefficient of variations of intra- and inter-assay imprecision were <8.2%, lower limit of quantification of 140 ng/L, and assay linearity up to 210,000 ng/L.

### Statistical analysis

Statistical analyses were performed using GraphPad Prism 5 (version 5.03 for Windows). As vitamin D and desmosine values were not normally distributed, Mann-Whitney U test was used to compare means between two groups, and Kruskal-Wallis test was used to compare means between multiple groups. Correlations were assessed over log-transformed data using a Pearson coefficient. A p-value of < 0.05 was used as the threshold for statistical significance.

## Results

### Vitamin D and disease outcome

The cohort included 135 patients whose baseline characteristics are extensively described elsewhere [2], and summarized in table 1. For 133 patients sufficient EDTA plasma was available to measure 25(OH)D levels during admission.

**Table 1:**
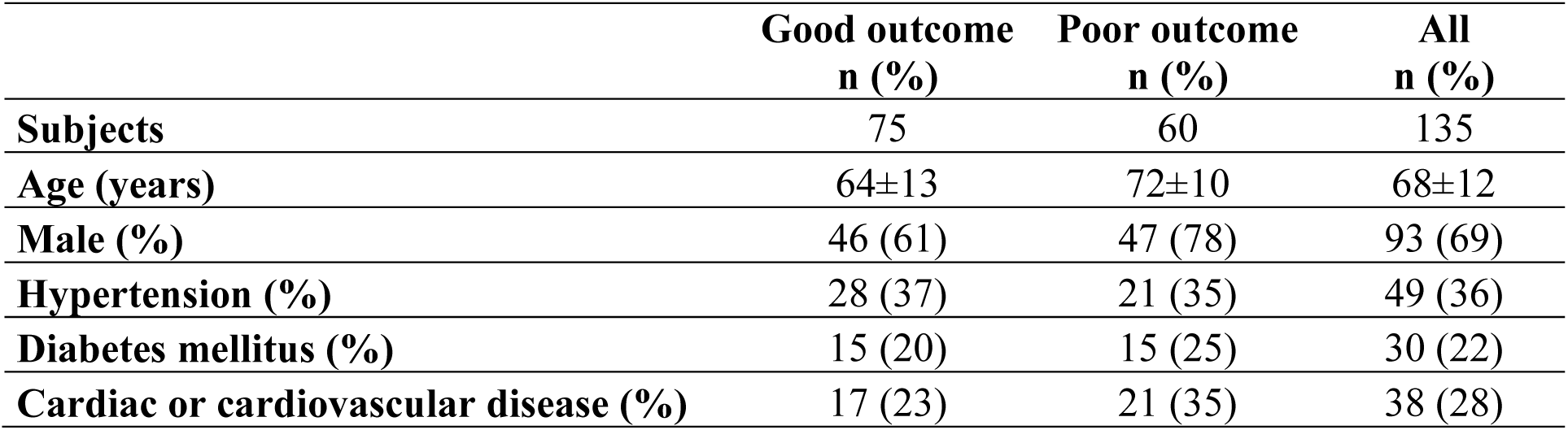
Baseline characteristics of COVID-19 patient cohort (n=135). Patient outcome was defined as ‘good’, i.e. survival without the need for invasive ventilation, or ‘poor’ i.e. the need for invasive ventilation and/or death.

25(OH)D levels were not significantly different between COVID-19 patients with good (n=75) and poor outcomes (n=58) (median 45.0 nmol/L interquartile range 26.7-67.9 nmol/L vs. median 37.7 nmol/L interquartile range 26.9-63.2 nmol/L, p=0.85), figure 1 and table 2.

**Table 2:** Outcomes of hospitalized COVID-19 patients (n=133) based on vitamin D status. Patient outcome was defined as ‘good’, i.e. survival without the need for invasive ventilation, or ‘poor’ i.e. the need for invasive ventilation and/or death.

| **25(OH) vitamin D** | **Good outcome**<br>**n (%)** | **Poor outcome**<br>**n (%)** | **Total**<br>**n (% of total)** | **25(OH)D**<br>**Median, range** |
| --- | --- | --- | --- | --- |
| <25 nmol/L | 13 (56) | 10 (43) | 23 (17) | 19.1, 5.80-24.6 |
| 25-50 nmol/L | 33 (58) | 24 (42) | 57 (43) | 30.9, 25.0-49.9 |
| >50 nmol/L | 29 (55) | 24 (45) | 53 (40) | 70.2, 50.1-115 |

**Figure 1:**
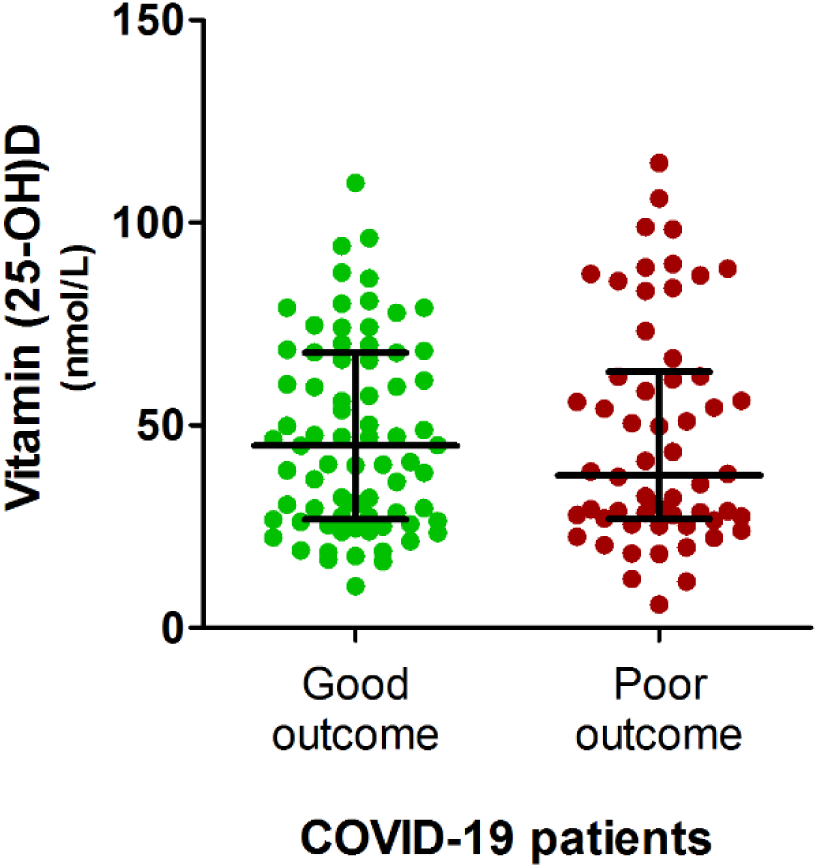
Vitamin D levels in hospitalized COVID-19 patients. 25(OH) vitamin D was measured in plasma from 133 patients. Patient outcome was defined as ‘good’, i.e. survival without the need for invasive ventilation, or ‘poor’ i.e. the need for invasive ventilation and/or death.

### Vitamin D and elastic fiber degradation

Desmosine was successfully measured in 125 patients. As plasma desmosine is strongly dialyzed (R. Janssen, unpublished data), three patients receiving dialysis at baseline were excluded from this analysis. Patients were separated into three categories based on vitamin D status: 1) vitamin D deficient (plasma levels lower than 25 nmol/L), 2) vitamin D insufficient (plasma levels between 25 and 50 nmol/L), or 3) vitamin D sufficient (plasma levels greater than 50 nmol/L). There was significant difference between desmosine levels between the groups (Kruskal-Wallis test p=0.019), with significantly lower desmosine in patients with 25(OH)D between 25-50 nmol/L compared to patients with 25(OH)D >50 nmol/L, figure 2A. There was no significant correlation between plasma desmosine and 25(OH)D levels (Pearson r=0.11, p=0.22).

**Figure 2:**
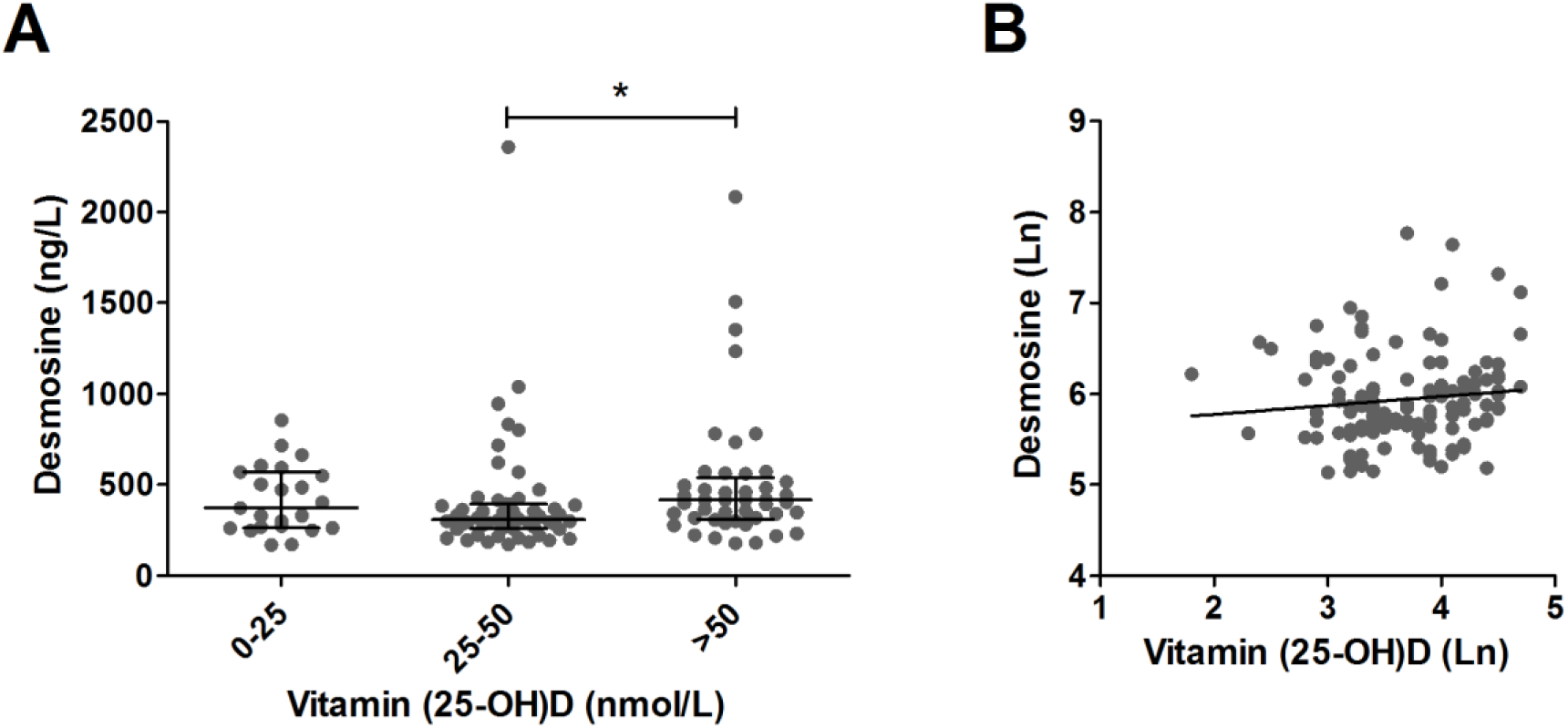
Effect of 25(OH) vitamin D status on elastic fiber degradation. (A) 25(OH) vitamin D and desmosine as a marker of elastic fiber degradation were measured in 122 COVID-19 patients. Dialysis patients were excluded from the analysis. Patients were divided based on vitamin D status. Lines and error bars represent median and interquartile range. **(B)** Correlation between desmosine and 25(OH) vitamin D, both log transformed. Line shows the result of a linear regression.

## Discussion

Here we investigated the role of vitamin D in patients hospitalized with COVID-19. We did not find a link between vitamin D deficiency and poor disease outcome, as we previously shown for vitamin K in the same cohort [2]. Elastic fiber degradation was accelerated in patients who were vitamin D sufficient compared to those with mild insufficiency.

The anti-inflammatory and antiviral potential of vitamin D [19], coupled with data from population-based and intervention studies [11], created a rationale to assess its protective effects against SARS-CoV-2 infections. A meta-analysis conducted before the COVID-19 pandemic demonstrated that daily and weekly administration of vitamin D reduces the risk of acute respiratory tract infections [11]. Previous vitamin D research in COVID-19 has largely been focused on observational studies that evaluated the infection rates in various populations [12, 13], as a measure of disease susceptibility. However, here we hypothesized that vitamin D status might also have effects on disease severity and outcome, as we had previously observed for vitamin K.

Approximately half of the patients in our cohort had vitamin D insufficiency or deficiency, comparable to the general incidence in The Netherlands [20]. In this cohort, vitamin D status was not associated with the need for invasive ventilation and/or death. In contrast with our findings, Panagiotou *et al*. recently found a higher incidence of vitamin D deficiency among British COVID-19 patients admitted to the intensive care [8]. However, like in our study, they did not find a difference in mean plasma 25(OH)D concentrations between the groups or an association with mortality.

In order to further evaluate a link between vitamin D status and pathology, we correlated 25(OH)D levels with the rate of elastic fiber degradation. Proteolytic activity and elastic fiber degradation are enhanced in severe COVID-19, both associating with poor disease outcome [2, 21]. We used plasma desmosine levels as a biomarker for tissue degradation, as it is an amino acid that only occurs in mature, cross-linked elastic fibers [22]. Its presence in the circulation therefore solely reflects elastolysis. We did not find a favorable decelerating effect of higher 25(OH)D levels on elastic fiber degradation, like we previously found for vitamin K [2]. Instead we showed that elastic fiber degradation was significantly lower in patients with a vitamin D insufficiency compared to those who were vitamin D sufficient. Patients with a more severe vitamin D deficiency had desmosine levels comparable to vitamin D sufficient persons.

We theorize that vitamin D might be like a double-edged sword in the pathogenesis of COVID-19, with both potential positive and negative effects. On the one hand, it might dampen deleterious inflammatory responses, reducing the cytokine storm [23]. On the other hand, vitamin D has a role in calcium metabolism and produces an increase in serum calcium [24]. Elastic fibers have high affinity for calcium [25]. Calcification is a pathological process that stimulates elastic fiber degradation and *vice versa* [26, 27]. This may explain an accelerating effect of vitamin D on the rate of elastic fiber degradation. Whereas systemic calcium is under strict homeostatic control, calcium binding to elastic fibers is expected to last after correction of circulating calcium levels [28]. During COVID-19, inflammatory infiltrates and proteases damage pulmonary elastic fibers [2]. These partially degraded fibers become more sensitive to calcium-ion binding [27], which in turn causes further degradation [29]. Through these mechanisms, vitamin D could both reduce inflammation by dampening cytokine response but increase proteolysis through calcification.

It can also be hypothesized that high-dose vitamin D supplementation during COVID-19 puts an excessive burden on already depleted vitamin K stores with subsequent deleterious consequences for elastic fibers [14, 15]. Evidence from animal models and other diseases supports this theory. In rats, administration of high doses of vitamin D induced rapid and severe calcification of the lungs and upregulation of pulmonary vitamin K-dependent MGP [15]. In kidney transplant patients, a population in which vascular calcifications are a major contributor to morbidity and mortality, vitamin D supplementation actually increased mortality in vitamin K deficient patients [30]. This is unsurprising considering the importance of vitamin K-activated MGP in protecting against elastic fiber calcification and degradation [15, 31]. Further combined analysis of vitamin D and vitamin K status, together with key markers of inflammation, protease activity and elastic fiber degradation during COVID-19 should shed light on the complex interplay between these factors.

A limitation of the current study is that it was conducted in one region in a single month, while vitamin D levels are known to vary strongly between populations and throughout the year. As such it will be important to validate these results in other cohorts. Yet, there was significant variability in vitamin D status between our patients, which strengthens our findings.

A small open-label pilot study recently suggested a decrease in ICU admissions after vitamin D administration [32], but larger controlled studies are needed to determine the role of supplementation in COVID-19. Regardless, considering the importance of vitamin K in protecting against the deleterious effects of calcium and the known interactions between vitamin D and vitamin K in calcium metabolism, it would be prudent to include investigation of vitamin K supplementation in any future clinical trial.

## Data Availability

Data will be made available upon reasonable request.

